# Awareness, Perception and Practice of COVID 19 Prevention among Residents of a State in the South-South Region of Nigeria: Implications for Public Health Control Efforts

**DOI:** 10.1101/2020.10.11.20210864

**Authors:** Golden Owhonda, Omosivie Maduka, Ifeoma Nwadiuto, Charles Tobin-West, Esther Azi, Chibianotu Ojimah, Datonye Alasia, Ayo-Maria Olofinuka, Vetty Agala, John Nwolim Paul, Doris Nria, Chinenye Okafor, Ifeoma Ndekwu, Chikezie Opara, Chris Newsom

**Affiliations:** Department of Public Health and Disease Control, Rivers State Ministry of Health; Department of Preventive and Social Medicine, University of Port Harcourt; Department of Community Medicine, Rivers State University; World Health Organization, Rivers State Field Office; Department of Internal Medicine, University of Port Harcourt; Rivers State Hospital Management Board; Stakeholder Democracy Network; Rivers State Public Health Emergency Operations Centre

**Keywords:** Covid-19 prevention, knowledge, attitude, practice, public health control

## Abstract

**Background:** This research explored awareness, perception, and practice of COVID 19 prevention among residents of communities in all the local government areas (districts) in Rivers State during the early stages of the pandemic response.

**Design:** This was a descriptive cross-sectional survey which employed an interviewer-administered four-page questionnaire built into the Open Data Kit application for android phones. Knowledge and practice scores were computed by scoring every correct response/action as 1 and wrong responses as 0. Knowledge was graded as excellent for scores of ≥80%, good for scores of 50-79% and poor for scores of <50%. Respondents who washed all critical parts of the hand were categorized as having correct handwashing practice.

**Setting:** Rivers State in the South-South region of Nigeria had recorded over 2000 cases of COVID 19 as of 18^th^ August 2020, ranking 5^th^ among the high burden states in Nigeria. As with any epidemic of an infectious nature, panic, fear, and misconceptions are rife. Risk communication utilizes multi-faceted activities geared towards facilitating correct and consistent knowledge and prevention practice.

**Participants:** Study involved 1,294 adult community residents in the 23 districts of the state.

**Results:** The respondents were aged between 18 and 80 years with average age of 39.6 years (SD = 11.9 years). A total of 710 (54.9%) were male, 476 (36.8%) were unemployed with 685 (52.9%) having secondary education. Almost all respondents 1,271 (98.2%) had heard about COVID 19. The three most common sources of information about COVID 19 were radio jingles 1102 (86.7%), television adverts 940 (74.0%) and announcements in Church 612 (48.2%). Overall, 608 (47.0%) of the respondents had poor knowledge of COVID 19. About 1167 (90.2%) of the respondents who were aware of COVID 19 acknowledged that COVID 19 is a problem in the state while 443 (34.9%) respondents believed they were unlikely contract the virus. Only 505 (39.0%) of the respondents washed all critical parts of the hand correctly.

**Conclusions:** Risk communication interventions during pandemics need to be based on an understanding of the gaps in knowledge, attitude, perceptions, and practice. Broadcast media has a pivotal role to play in risk communication for behaviour change for the control of current and future epidemics in this population.

## Introduction

The novelle coronavirus disease (COVID 19) caused by the severe acute respiratory syndrome coronavirus 2 (SARS – Cov 2) was first found in Wuhan, China in 2019. ^1^ The disease spread quickly in epic proportions to over 26 countries within eight weeks, prompting the World Health Organization (WHO) to declare it a pandemic on the 11^th^ of March 2020.^2^ The pandemic has had its toll on virtually every country in the world, including Nigeria, which recorded its 1^st^ case in February 2020 and has since gone on to record over 50 000 cases as at 18^th^ of August 2020.^3^ Rivers State is in the South-South region and a major economic city known for crude oil exploration in the country has had its share of the pandemic, with close to over 2000 cases as at 18^th^ of August 2020, following its index case identified on the 25^th^ of March 2020 ranking 5^th^ among the high burden states in Nigeria.^4^

As with any epidemic of an infectious nature, panic, fear, and misconceptions are rife. This is more so because COVID 19 does not presently have a vaccine or a specific cure.^1^ Coronavirus disease is known to be highly infectious but with a low National case fatality rate of 2.1% compared to other epidemic diseases like Ebola virus disease (42.1%) and Lassa Fever disease (20.9%) that have hit the country.^5^ A similar picture of misinformation, poor knowledge on disease outbreak and negative behaviours was observed during the Ebola epidemic of 2014, which led to a lot of harmful practices carried out by people to remain safe.^6,7^ A study done in China in the early stages of the pandemic showed good knowledge and practice of preventive measures against COVID 19 with 98% of respondents wearing a facemask when going out. ^8^ However, in Nigeria, while most people identified radio as their main source of information on COVID 19, they attributed the disease to one affecting only the affluent and could not be bothered with the practice of preventive measures in the face of economic hardship during the lockdown.^9,10^

The risk communication pillar of the public health emergency operations centre (PHEOC) is responding to the pandemic through multi-faceted activities relating to community mobilization, mass media, training of message multipliers, information education and communication (IEC) materials etc. These activities are geared towards facilitating correct and consistent information from experts to communities at risk to enable them to adopt behaviours to prevent and control COVID 19.^12^ The efforts at flattening the pandemic curve in the South-South region will ultimately depend on the willingness of people to adopt and maintain public health preventive health practices as advocated during community engagements.

It is therefore critical that a risk assessment is carried out to explore awareness, perception, and practice of COVID 19 prevention among residents of communities in Rivers State. This would advise risk communication activities to facilitate desirable behaviour change for COVID 19 and address rumours and misconceptions gotten as feedback from the survey.

## Methods

This was a descriptive cross-sectional survey which took place among community residents in all 23 Local Government Areas (LGAs) of Rivers State between 18^th^ May to 10^th^ June 2020. Data was collected by disease surveillance and notifications officers (DSNOs) who are familiar with the terrain and in each LGA and are trained in data collection techniques. A sample size of 1,186 adult respondents was calculated based on the sample size formula for single proportions where the prevalence of good knowledge about COVID 19 was set at 50% (since there is no previously established prevalence), degree of accuracy set at 3% and 10% non-response rate.

The data collection tool was an interviewer-administered four-page questionnaire built into the Open Data Kit application for android phones with GPS tracking. A training was done for the DSNOs on the objectives and tools for the study. Data was exported to Microsoft Excel and analysed with SPSS version 23.

Major outcome variables studied were knowledge of COVID 19, attitude towards COVID 19 and practice of COVID 19 prevention methods. Knowledge score was computed by scoring every correct answer to the knowledge questions as 1 and wrong answers as 0. The total knowledge score was computed, and the average score calculated by dividing it by the total number of knowledge question. Knowledge was then graded as excellent for scores of ≥80%, good for scores of 50-79% and poor for scores of <50%. To compute for handwashing practice, respondents who washed all critical parts of the hand were categorized as having correct handwashing practice, while those who did not were classified as having incorrect handwashing practice. Descriptive statistics was done, and results presented in tables.

## Results

A total of 1,294 persons consented to participate in the study. The respondents aged between 18 and 80 years with 422 (32.6%) aged between 35 and 44 years. The average age was 39.6 years with a standard deviation of 11.9 years. More than half of the respondents 710 (54.9%) were male, more than a third were unemployed 476 (36.8%) with more than half having secondary education as their highest level of education 685 (52.9%). ***Table 1***

**Table 1:** Socio-demographics characteristics of Study participants

| Variables | Frequency<br>(n=1294) | Percentage<br>(%) |
| --- | --- | --- |
| **Age Group** |  |  |
| Less than 25 years | 110 | 8.5 |
| 25-34 years | 347 | 26.8 |
| 35-44 years | 422 | 32.6 |
| 45-54 years | 260 | 20.1 |
| 55-64 years | 120 | 9.3 |
| 65 years and above | 35 | 2.7 |
| **Mean±SD** | 39.6±11.9 years |  |
| Median | 38 years |  |
| Mode | 36 years |  |
| Range | 18-80 years |  |
| **Gender** |  |  |
| Male | 710 | 54.9 |
| Female | 584 | 45.1 |
| **Occupation** |  |  |
| Senior public servants; professional; manager; large scale traders; businessman; contractors | 235 | 18.2 |
| Intermediate grade public servants and senior schoolteachers | 117 | 9.0 |
| Junior schoolteachers; drivers; artisans | 107 | 8.3 |
| Petty traders; labourers; messengers and similar | 359 | 27.7 |
| Unemployed | 476 | 36.8 |
| **Religion** |  |  |
| Christian | 1246 | 96.3 |
| Islam | 19 | 1.5 |
| Traditional | 23 | 1.8 |
| Others | 6 | 0.5 |
| **Highest Level of Education** |  |  |
| No formal education | 38 | 2.9 |
| Primary education | 120 | 9.3 |
| Secondary education | 685 | 52.9 |
| Tertiary education | 451 | 34.9 |

Almost all respondents 1,271 (98.2%) had heard about COVID 19. The three most common sources of information about COVID 19 were radio jingles 1102 (86.7%), television adverts 940 (74.0%) and announcements in Church 612 (48.2%). Only 441 (34.7%) of those who had heard of COVID 19 were aware that the disease was caused by a virus, 299 (23.5%) wrongly associated COVID 19 to causes other than a virus, while 531 (41.8%) reported that they do not know the cause of COVID 19. ***Table 2***.

**Table 2:** Awareness of COVID 19 in the Rivers State

| **Heard of COVID 19 (n=1294)** | **Frequency (n=1271)** | **Percentage (%)** |
| --- | --- | --- |
| No | 23 | 1.8 |
| Yes | 1271 | 98.2 |
| **Source of information about COVID 19 (Multiple responses)** |  |  |
| Radio | 1102 | 86.7 |
| Television | 940 | 74.0 |
| Church | 612 | 48.2 |
| Social Media (Facebook, Twitter, WhatsApp) | 607 | 47.8 |
| GSM/SMS | 568 | 44.7 |
| Peers/Friends | 523 | 41.1 |
| Family Member | 512 | 40.3 |
| Town Announcer | 501 | 39.4 |
| Health Facility | 490 | 38.6 |
| Newspaper | 410 | 32.3 |
| Flyer | 400 | 31.5 |
| Health Educator | 371 | 29.2 |
| Neighbourhood | 340 | 26.8 |
| Internet Sites | 336 | 26.4 |
| Market | 315 | 24.8 |
| Journal | 222 | 17.5 |
| Mosque | 76 | 5.9 |
| Others | 2 | 0.2 |
| **Causes of COVID 19** |  |  |
| Virus | 441 | 34.7 |
| Causes other than a virus | 299 | 23.5 |
| Do not know | 531 | 41.8 |

About 1,112 (87.5%) of the respondents who were aware of COVID 19 reported that the virus spreads through contact with a person who is sick of coronavirus disease, 743 (58.5%) reported that it spreads through contact with contaminated surfaces, beddings, or clothing of a person who is sick of coronavirus disease, 727 (57.2%) reported that it spreads through hugging and kissing, 718 (56.5%) reported that it spreads through touching blood, urine, stool or saliva from a person who is sick of coronavirus disease, and 698 (54.9%) reported that it spreads through participation in burial rites of a person who has died from coronavirus disease. Only 36 (2.8%) of the respondents reported that they do not know how COVID 19 spreads.

The three most frequent symptoms of COVID 19 identified by the respondents were fever 1,187 (93.4%) cough 1,134 (89.2%) and difficulty in breathing 1,010 (79.5%). A total of 834 (66.4%) of respondents were aware that the signs and symptoms of COVID 19 manifest in an infected person between 2 to 14 days after contracting the virus, 967 (76.1%) were aware that there is no specific drug treatment for COVID 19, and 1006 (70.2%) were aware that there was no specific vaccine for COVID 19. Overall, 608 (47.0%) of the respondents had poor knowledge of COVID 19, 587 (45.4%) had good knowledge, while only 99 (7.7%) had excellent knowledge about COVID 19. ***Table 3***

**Table 3:** Respondents knowledge of COVID 19 across the 23 LGAs of Rivers State

|  | Frequency<br>(n=1271) | Percentage<br>(%) |
| --- | --- | --- |
| **How COVID 19 spreads (Multiple responses)** |  |  |
| Contact with a person who is sick of coronavirus disease | 1112 | 87.5 |
| Contact with contaminated surfaces, beddings, clothing of a person who is sick of coronavirus disease | 743 | 58.5 |
| Hugging, kissing | 727 | 57.2 |
| Touching blood, urine, stool, or saliva from a person who is sick of coronavirus disease | 718 | 56.5 |
| Participating in burial rites of a person who has died from coronavirus disease | 698 | 54.9 |
| Sharing sharp objects such as razors, needles e.t.c with a person who has coronavirus disease | 562 | 44.2 |
| Through the air | 467 | 36.7 |
| Through sexual intercourse | 331 | 26.0 |
| From infected animals to man | 247 | 19.4 |
| I do not know | 36 | 2.8 |
| Others | 23 | 1.8 |
| **Signs and symptoms of COVID 19 (Multiple responses)** |  |  |
| Fever | 1187 | 93.4 |
| Cough | 1134 | 89.2 |
| Difficulty in breathing | 1010 | 79.5 |
| Sore throat | 734 | 57.7 |
| Weakness | 587 | 46.2 |
| Headache | 584 | 45.9 |
| A general feeling of unwell | 442 | 34.8 |
| Body pain | 243 | 19.1 |
| Diarrhoea | 165 | 13.0 |
| Vomiting | 158 | 12.4 |
| Rash on the body | 107 | 8.4 |
| Sneezing | 34 | 2.7 |
| Others | 2 | 0.2 |
| **When signs and symptoms of COVID 19 manifest** |  |  |
| In less than 2 days | 39 | 3.1 |
| Between 2 and 14 days | 844 | 66.4 |
| After 14 days | 142 | 11.2 |
| I do not know | 246 | 19.4 |
| **There is specific drug treatment for COVID 19** |  |  |
| No | 967 | 76.1 |
| Yes | 47 | 3.7 |
| I do not know | 257 | 20.2 |
| **There is a specific vaccine for COVID 19** |  |  |
| No | 1006 | 79.2 |
| Yes | 15 | 1.2 |
| I do not know | 250 | 19.7 |
| **Overall knowledge score** |  |  |
| Poor knowledge | 608 | 47.7 |
| Good knowledge | 567 | 45.5 |
| Excellent knowledge | 99 | 7.8 |

About 1167 (90.2%) of the respondents who are were aware of COVID 19 acknowledged that COVID 19 is a problem in the state because; It has no cure 817 (70.0%), it’s highly infectious 797 (68.3%), It’s a deadly disease 788 (67.5%), and creates a lot of panic 779 (66.8). Those who did not consider COVID 19 a problem reported that it is just being exaggerated 64 (50.4%), that people just want to make money with coronavirus intervention 52 (40.9%), and that they do not believe that there are cases of coronavirus 47 (37.0%).

Four hundred and forty-three (34.9%) of the respondents believed they cannot contract the virus, while 801 (63.0%) acknowledged that the government is doing enough to contain the virus. Most respondents would go to the hospital 905 (71.2%) or call the coronavirus help number 868 (68.3%) if they develop signs and symptoms of COVID 19. A total of 743 (58.5%) of the respondents had heard of the coronavirus helplines but only 116 (15.6%) of them could make it readily available when requested for. Also, 522 (41.1%) had heard of the NCDC coronavirus information website/social media account, but only 53 (10.2%) of them could make it readily available when requested for. A total of 833 (65.5%) of the respondents claimed they were comfortable living or working with a person treated for COVID 19. ***Table 4***

**Table 4:**
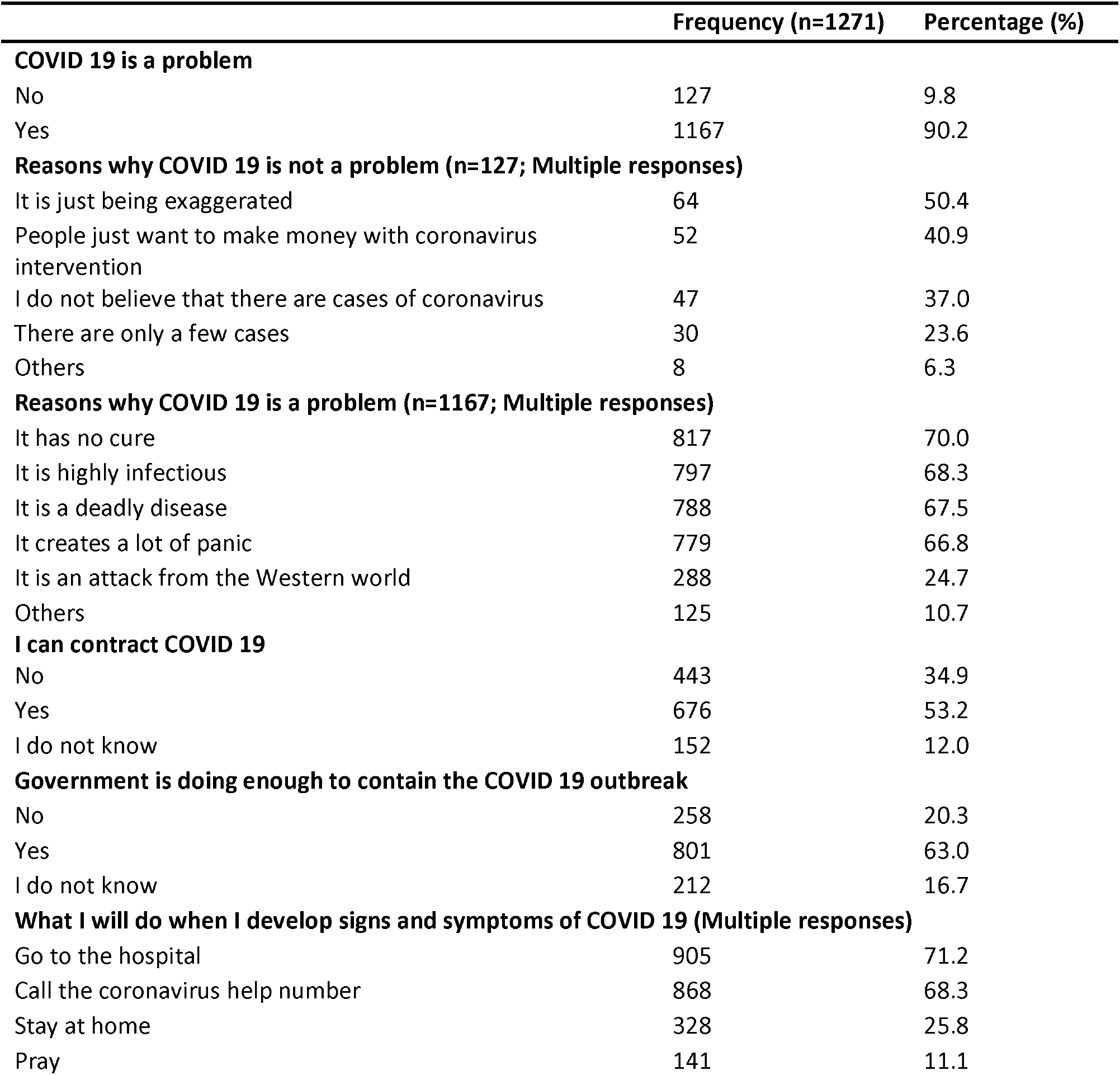
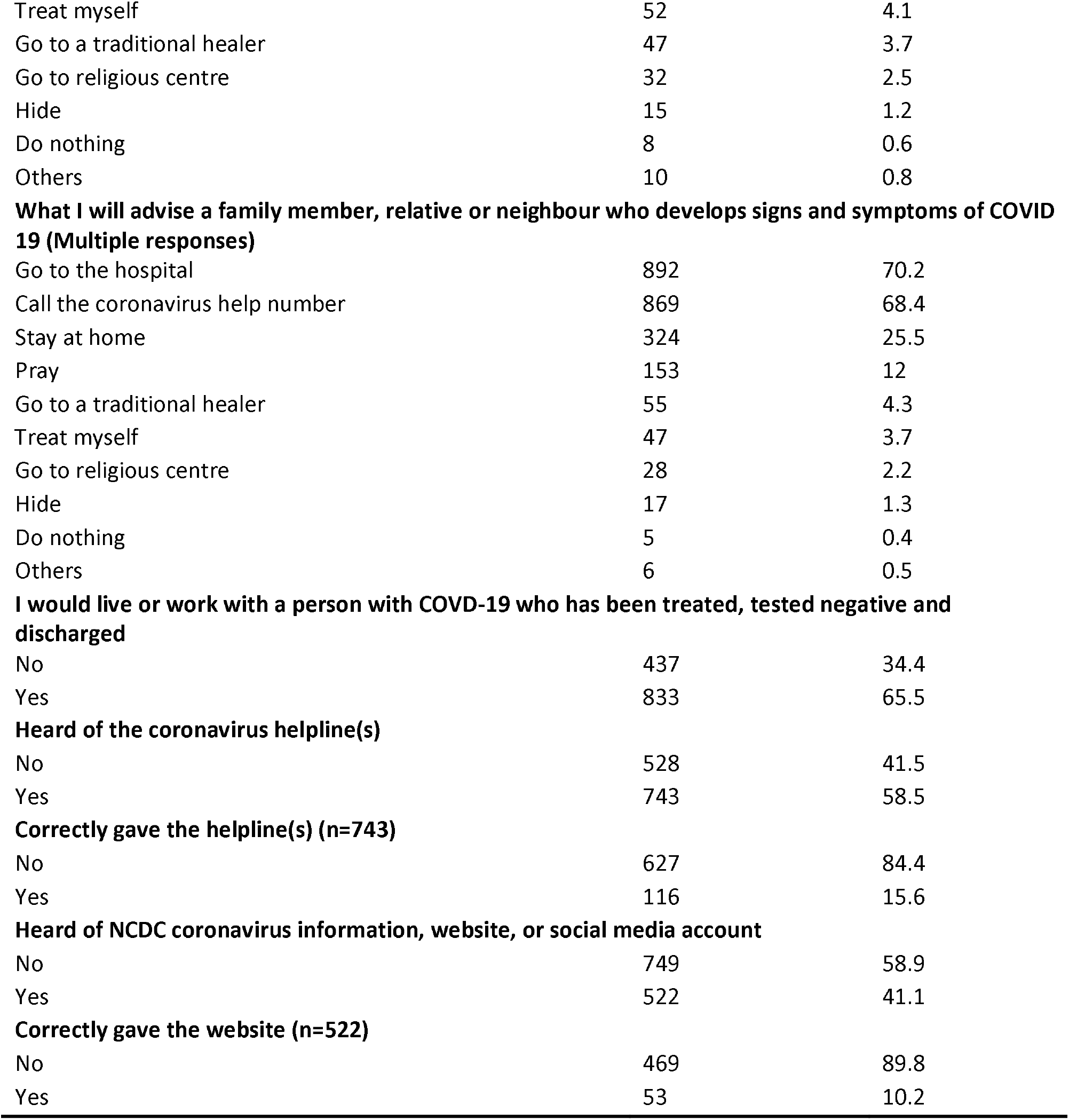
Attitude of respondents towards the COVID 19 outbreak in Rivers State

Among the respondents who had heard of COVID 19, 1150 (90.5%) reported that they prevent COVID 19 by practising regular handwashing with soap and water, 1081 (85.1%) by maintaining physical distance, 857 (67.4%) by regular use of hand sanitizer, 815 (64.1%) by avoiding crowded places/events. During the demonstration of handwashing, only 505 (39.0%) of the respondents washed all critical parts of the hand correctly. ***Table 5***

**Table 5:** Practice for prevention of COVID 19 transmission

|  | Frequency<br>(n=1271) | Percentage<br>(%) |
| --- | --- | --- |
| **How you prevent yourself from getting COVID 19 (Multiple responses)** |  |  |
| Regular handwashing with soap and water | 1150 | 90.5 |
| By keeping your distance from any person with suspected coronavirus disease | 1081 | 85.1 |
| Regular use of hand sanitizer | 857 | 67.4 |
| By avoiding crowded places/events | 815 | 64.1 |
| By staying at home | 681 | 53.6 |
| Regular handwashing with water only | 105 | 8.3 |
| Regular gargling/drinking of water | 66 | 5.2 |
| By drinking concoctions (lime, ginger, garlic, etc) | 65 | 5.1 |
| Taking chloroquine | 59 | 4.6 |
| Going for special prayers | 49 | 3.9 |
| Eating bitter kola | 42 | 3.3 |
| Avoid eating bushmeat | 35 | 2.8 |
| Bathing with saltwater | 25 | 2.0 |
| Others | 16 | 1.3 |
| I do not know | 14 | 1.1 |
| **Washed all critical parts of the hand (n=1294)** |  |  |
| No | 789 | 61.0 |
| Yes | 505 | 39.0 |

## Discussion

It was heart-warming that awareness about COVID 19 infection among community-based residents in the State was high, a few months after the index case of the disease was recorded. This was an affirmation of broadcast media (radio and television) as the most viable channel for risk communication within the State. Radio and Television as potent sources of information for the people and stand out as channels for epidemic response efforts. They should therefore be engaged early and effectively. The high level of awareness of COVID 19 found in this study, was similar to that obtained in China early in the outbreak,^8^ but contrasted significantly from other studies in Nigeria, where COVID 19 awareness remained poor several weeks after the pandemic.^9,10^ This observed awareness was closely followed by good knowledge scores on COVID 19 transmission, signs, and symptoms of the disease such as fever, cough and breathing difficulties in more than half of study respondents. This is also commendable because knowledge of modes of disease transmission and signs and symptoms of a disease is the first proactive step in understanding the measures of disease prevention and control, otherwise known as primary prevention. Nevertheless, the good knowledge was not observed for slightly less than half of respondents and did not sufficiently translate to good prevention practices among the people. This was because while regular hand washing was reported to have been practised by most respondents, only a third of the respondents satisfactorily demonstrated all handwashing steps. This supports the assertion that knowledge does not always automatically translate into good practice without going through the structured stages of declarative, procedural, and autonomous processes involved in the transition from knowledge to practice.^11^ The finding has huge implications for the prevention and control of the epidemic in the state and therefore cannot be glossed over by the Risk Communication Pillar of the State Public Health Emergency Response. It suggests a paradigm shift in the risk communication process in reprogramming its communication strategies to include skills based packages, in order to make any significant, meaningful impacts in healthful behaviour change among the people concerning the COVID 19 virus transmission truncation in the state.

Also, found was the disturbing fact that a third of participants responded with a stigmatizing attitude towards persons who have been treated for COVID 19. This was not peculiar to our study alone as other researchers have reported similar occurrences in their environments.^12,13^ The fact that the disease is new, with many unknowns and the fact people are often afraid of the unknown, evokes the fear factor among individuals, including community dwellers. It is therefore understandable why there is confusion, anxiety, and fear among the public. Unfortunately, however, these factors are also fuelling harmful stereotypes and labels of individuals, which in turn drive discrimination and loss of status because of a perceived link with the disease.^12^ The aftermath of which can drive the epidemic underground by making people hide the illness to avoid discrimination, prevent people from seeking health care immediately and discourage them from adopting healthy behaviours. All of these can result in more severe health problems in the State by increasing the difficulties of controlling the disease outbreak.

Another salient issue that might have promoted stigma and discrimination was the unprofessional manner the print and broadcast media portrayed the disease at inception, as a highly infectious and deadly disease. And this created a lot of panic in the minds of the people as many persons developed the perception that the disease as a huge problem. This thus created a paradox in the COVID 19 risk communication in the State and further underscores the importance of periodic media engagement and capacity building in ethical reporting and accurate information dissemination since the media are critical partners in disease outbreak response.^14^

This study brings to the fore the relevance of adopting skills-based approaches in health risk communication and the timely and effective media engagement on ethical reporting of health matters during disease outbreaks to mitigate stereotyping, stigmatization and discrimination, all which have severe implications for epidemic prevention and control.^15^ It is therefore recommended that the Risk Communication Pillar of the Public Health Emergency Operations Centre for COVID 19 in Rivers State reprogramme its communication strategy to include sufficient practice elements that demonstrate the correct COVID 19 prevention methods like hand hygiene, social distancing and appropriate use of face masks.

The study was of a cross-sectional design which was based on self-reporting and therefore subject to information bias, whereby respondents might choose not to give information as required. However, the large sample size and spread of respondents across all the LGAs of the state make this a relevant piece of research.

## Conclusion

Risk communication interventions during pandemics need to be based on an understanding of the gaps in knowledge, attitudes, perceptions, and practice. Patronised communication channels such as broadcast media should be maximally utilized to foster behaviour change communication for the control of current and future epidemics.

## Data Availability

Data for this research is available on request

## Acknowledgements

1. Disease Surveillance and Notification Officers of the Rivers State Primary Health Care Management Board for their role in data collection
2. Stakeholder Democracy Network for funding the survey
3. Technical and Administrative Staff of the Rivers State Public Health Emergency Operations Centre for the key roles they play in the pandemic response

